# Smoking, all-cause, and cause-specific mortality in individuals with diabetes in Mexico: an analysis of the Mexico City Prospective Study

**DOI:** 10.1101/2023.09.19.23295818

**Authors:** Daniel Ramírez-García, Carlos A. Fermín-Martínez, Paulina Sánchez-Castro, Alejandra Núñez-Luna, Martín Roberto Basile-Alvarez, Luisa Fernández-Chirino, Arsenio Vargas-Vázquez, Juan Pablo Díaz-Sánchez, Ashuin Kammar-García, Paloma Almeda-Valdés, Jaime Berumen-Campos, Pablo Kuri-Morales, Roberto Tapia-Conyer, Jesus Alegre-Díaz, Jacqueline A. Seiglie, Neftali Eduardo Antonio-Villa, Omar Yaxmehen Bello-Chavolla

## Abstract

**BACKGROUND:** Evidence from low– and middle-income countries regarding the effect of smoking in people with diabetes is lacking. Here, we report the association of smoking with mortality in a large cohort of Mexican adults with diabetes.

**METHODS:** Participants with diabetes mellitus (self-reported medical diagnosis, use of antidiabetic medications or HbA1c ≥6.5%) aged 35-74 years when recruited into the Mexico City Prospective Study were included. Cox regression confounder-adjusted mortality rate ratios (RRs) associated with baseline smoking status were estimated.

**FINDINGS:** Among 15,975 women and 8225 men aged 35-74 years with diabetes but no other comorbidities at recruitment, 2498 (16%) women and 2875 (35%) men reported former smoking and 2753 (17%) women, and 3796 (46%) men reported current smoking. Smoking less than daily was common: 39.4% of female current smokers and 30.7% of male current smokers. During a median of 17 years of follow-up there were 5087 deaths at ages 35-74 years. Compared with never smoking, the all-cause mortality RR was 1.08 (95%CI 1.01–1.17) for former smoking, 1.09 (95%CI 0.99–1.20) for non-daily smoking, 1.06 (95%CI 0.96–1.16) for smoking <10 cigarettes per day (median during follow-up 4 cigarettes/day) and 1.28 (95% CI 1.14–1.43) for smoking ≥10 cigarettes/day (median during follow-up 15 cigarettes/day). Excess mortality among current daily smokers was greatest for COPD, lung cancer, cardiovascular diseases, and acute diabetic complications.

**INTERPRETATION:** In this cohort of Mexican adults with diabetes, low-intensity daily smoking was associated with increased mortality, despite observing smoking patterns which are different from other populations. Of those smoking ≥10 cigarettes per day, over one fifth were killed by their habit. Quitting substantially reduced these risks.

**RESEARCH IN CONTEXT:** *Evidence before this study:* We conducted a search in PubMed to identify studies on smoking-related mortality in individuals with diabetes before September 1^st^, 2023, using the terms ((diabetes mellitus) OR (type 2 diabetes mellitus)) AND ((smoking) OR (tobacco)) AND ((mortality) OR (survival analysis) OR (death)). We identified a meta-analysis published in 2015 which analysed data from 1,132,700 participants with diabetes from 89 cohort studies. Among current and former smokers, the pooled adjusted relative risk for all-cause mortality was 1.55 (95% CI 1.46–1.64) and 1.19 (95% CI 1.11–1.28), respectively. Among current smokers, there was considerably higher risk of death due to cardiovascular disease, coronary heart disease, stroke, peripheral arterial disease, and heart failure. Most included studies were conducted in high income countries and only one was done in Latin America (Argentina, 1,730 participants). In Mexico, epidemiological studies related to smoking have been primarily descriptive in nature. A previous study of the MCPS that analysed all-cause smoking-related mortality among disease-free individuals found adjusted rate ratios among daily smokers of 1.17 (95% CI 1.10–1.25) for those using <10 cigarettes/day and 1.54 (95% CI 1.42–1.67) for those using ≥10 cigarettes/day. In this study, mortality risk in people with diabetes was not assessed separately. Instead, mortality risk estimates were obtained for several chronic diseases as a group. No other studies have analysed changes in smoking habits and smoking-related mortality in Mexican individuals with diabetes.

*Added value of this study:* To our knowledge, this is the first analysis of longitudinal data of Mexican adults with diabetes (n=24,200) followed over 17 years, to examine the association between smoking and all-cause and cause-specific mortality. Compared with never smokers with diabetes, former and daily smokers with diabetes had elevated mortality. Among daily smokers there was higher risk of death due to cardiovascular disease, acute diabetes complications, lung cancer and COPD. Our results are generally consistent with previous analyses that have found smoking as a significant risk factor for higher mortality in individuals with diabetes. Our estimates, however, are comparably lower for all-cause mortality, possibly related to distinct smoking habits specific to the Mexican population with diabetes.

*Implications of all available evidence:* Our results confirm that smoking is a significant risk factor for all-cause and cause-specific mortality among Mexican adults with diabetes. Strategies to promote smoking cessation among individuals with diabetes should be encouraged, particularly given that tobacco use is a modifiable risk factor. This is especially meaningful in middle-to lower-income countries like Mexico, where diabetes prevalence is increasing and achieving glycaemic targets remains a challenge.

## INTRODUCTION

Diabetes mellitus is a significant public health problem, with over 530 million individuals living with the disease and around 6.7 million deaths attributable to diabetes worldwide in 2021^1^. In Mexico, diabetes prevalence has tripled in the last three decades^2,3^ and currently affects one in six Mexican adults^1^. Besides the microvascular complications associated with this condition, diabetes is a well-documented accelerant of cardiovascular disease (CVD), which is the leading cause of premature mortality in Mexico. While comprehensive risk factor management to reduce CVD risk in people with diabetes is an integral aspect of diabetes care^4^, addressing modifiable risk factors to reduce CVD burden remains a challenge in Mexico^2^. A key pervasive modifiable risk factor for diabetes and CVD incidence and mortality is tobacco smoking. Despite ratification of the WHO’s Framework on Tobacco Control (FCTC) in 2014, prevalence of current smoking among Mexican adults has remained relatively stable in the past decade, with nearly 1 in every 5 Mexican adults reporting current smoking in 2016^5^. While broader implementation of the FCTC is needed, characterizing mortality risk attributed to smoking in Mexican adults with diabetes could help inform tailored strategies for smoking cessation in this population.

The Mexico City Prospective Study (MCPS) is a large blood-based prospective study of 150,000 adults who were recruited between 1998 and 2004 and have been tracked for cause-specific mortality ever since. A previous analysis of the MCPS identified an increased risk of all-cause mortality among disease-free former and current smokers. Risk of all-cause mortality was increased even among adults who reported low-intensity daily smoking. However, participants with diabetes and other chronic diseases were excluded to minimize reverse causality^6^. Despite the important contribution of this analysis to the literature on smoking and mortality, smoking patterns in people with diabetes as well as smoking-related mortality in Mexican adults with diabetes remains unexplored. In this study, we aim to characterize patterns of tobacco use in individuals with diabetes and to estimate the effect of smoking on all-cause and cause-specific mortality, exploring the influence of diabetes-related factors (glycaemic control and disease duration) as potential modifiers of this association.

## METHODS

### Cohort characteristics

We analysed data from participants enrolled in the MCPS, a prospective, population-based cohort study with a baseline survey conducted from 1998 to 2004^7^. Households from two urban districts of Mexico City (Coyoacán and Iztapalapa) were visited, and every adult ≥35 years was invited to participate. In total, 159,755 individuals were recruited, and information was collected regarding age, sex, lifestyle habits, self-reported medical history, use of medications, and socio-demographic information including education, civil status, occupation, and monthly income. Clinical measurements (weight, height, and blood pressure) were obtained, and a non-fasting 10-mL blood sample was collected for subsequent analysis. From 2015-2019, a follow-up survey was conducted on 10,143 surviving participants, obtaining similar data regarding lifestyle habits, medical history, clinical measurements, and another blood sample as described previously^8^. Blood samples, both at baseline and resurvey examinations, were stored overnight in a central laboratory, separated into plasma and buffy coat, and then frozen to –80°C. Afterwards, samples were shipped to the University of Oxford for analysis and storage. Assays of HbA1c were performed from buffy coat samples in the Clinical Trial Service Unit and Epidemiological Studies Unit’s Wolfson laboratory, which has International Organization for Standardization (ISO)-17025 accreditation, as described previously^9^.The study protocol was approved by the corresponding ethics committees at the Mexican Ministry of Health, the Mexican National Council for Science and Technology, and the University of Oxford. All study participants provided written informed consent.

### Study population

We defined diabetes mellitus according to self-reported medical diagnosis of diabetes, use of diabetes medication (biguanides, sulfonylureas, insulin or other), or HbA1c ≥6.5%^10^. Diagnosed diabetes was defined as previous medical diagnosis of diabetes or use of glucose lowering medication and undiagnosed diabetes as HbA1c ≥6.5% in an individual without a previous diagnosis of diabetes. Given the heterogeneity of diabetes mellitus^11^, and to better account for diabetes-related variables, the primary analysis was restricted to individuals with previously diagnosed or undiagnosed diabetes mellitus, excluding participants without diabetes. To address the possibility of reverse causality in the association between smoking and all-cause mortality, we excluded participants who reported comorbidities other than diabetes at baseline, including ischaemic heart disease, pulmonary disease, chronic kidney disease, cirrhosis, or cancer. We also excluded individuals with missing covariates or with uncertain mortality linkage.

### Smoking related variables

We defined smoking status according to baseline interview information. Never smokers were classified as those who reported never smoking, former smokers as those who reported having smoked previously but not currently and current smokers as those having ever smoked and smoking currently (daily or non-daily). We classified smoking intensity into five categories (never smoker, ex-smoker, current non-daily smoker, current daily smoker <10 cigarettes per day, and current daily smoker ≥10 cigarettes per day), and age when started smoking into three categories (<15 years, 15-23 years, ≥24 years). In former smokers, no data was available regarding time since they stopped smoking; therefore, we were unable to explore this characteristic. We also analysed changes in smoking status between baseline interview and resurvey to explore changes in smoking behaviour over the study period. To do this, we matched surviving individuals who were included in the main mortality analysis at baseline and resurveyed during the 2015-2019 period. Then, we calculated the proportion of participants with a given smoking status at baseline and the proportion at resurvey.

### Mortality follow-up

Mortality follow-up of participants was done through electronic probabilistic linkage to death registries, with deaths tracked up to December 31^st^, 2020, and registered according to the International Classification of Diseases 10^th^ Revision (ICD-10). Cause-specific mortality was classified into several categories (cardiovascular, myocardial infarction, stroke, other vascular, acute diabetes complications [diabetic coma and ketoacidosis], lung cancer, non-lung cancer, COPD, other respiratory, renal, hepatobiliary, and gastric) in accordance with the underlying causes of death determined by study clinicians. The specific ICD-10 codes used for every cause and the number of deaths in each of them are provided in **Supplementary Table 1.**

### Statistical analyses

For descriptive analyses, we calculated mean and standard deviation in continuous variables (median and interquartile range for HbA1c, number of cigarettes per day and age started smoking), and proportions for categorical variables. To explore the association between smoking, all-cause and cause-specific mortality amongst individuals with diabetes, we fitted Cox proportional hazard regression models to estimate adjusted hazard ratios (HR) for former and current smokers using individuals with diabetes who were never smokers as the reference group with analyses restricted to premature mortality (deaths at ages 35-74 years). All models were stratified by sex and age at risk (5-year age groups)^12^ and adjusted for place of residence (Coyoacán or Iztapalapa), educational level (elementary, high school, university, other), body mass index (BMI), HbA1c, time since diabetes diagnosis, and alcohol consumption. We tested the proportional hazards assumption using Schoenfeld residuals. As sensitivity analyses, we fitted separate Cox models for males and females to further characterize whether there are different risk profiles by sex. We estimated attributable fraction with the formula (RR – 1) / RR. In the text, conventional 95% confidence intervals (CI) comparing two groups are used. In the figures, however, group specific 95% CIs, are shown for every RR (including the reference group); these were estimated using floating absolute risks to reflect the amount of information in each category^13^. All statistical analyses were conducted using R software version 4.2.1.

## RESULTS

### Smoking status and baseline characteristics of participants with diabetes

Among 159,755 participants recruited from 1998 to 2004, we identified 29,948 individuals with diabetes at baseline. Of these, 3,953 were excluded because they had comorbidities other than diabetes (ischemic heart disease, pulmonary disease, chronic kidney disease, cirrhosis, or cancer) and another 1,795 were excluded due to missing covariate or mortality data. In total, we included 24,200 participants with diabetes with complete data on the mortality analyses (**Supplementary** Figure 1). Among women, 10,724 (67.1%) reported being never smokers, 2,498 (15.6%) former smokers, and 2,753 (17.2%) current smokers at baseline, of whom 1,667 (60.6%) were daily smokers and 1,086 (39.4%) non-daily smokers (**Table 1**). Compared to female never smokers, female daily smokers were younger, had worse glycaemic control (especially those smoking <10 cig/d) and a slightly higher body mass index (BMI). Among men, there was a lower proportion of never smokers with 1,554 (18.89%), and higher proportions of former and current smokers, with 2,875 (34.95%) and 3,796 (46.2%), respectively. Among current smokers, 2,629 men (69.3%) were daily smokers and 1,167 (30.7%) were non-daily smokers. Compared to male never smokers, male daily smokers were younger, had worse glycaemic control, and the median age at which they started smoking was 17 years (IQR 15, 20). In women, the greatest proportion of participants were never smokers and the proportion of current smokers decreased with age. Cessation, however, was less common in this group with a consistent number of former smokers across all age cohorts (**Figure 1**). In men, on the other hand, there was a higher proportion of current smokers and the proportion of former smokers increased with age. The proportion of never smokers was relatively low regardless of age. Amongst current smokers, the prevalence of non-daily smoking was higher at younger ages for both men and women, low-intensity daily smoking (<10 cig/day) increased for older individuals irrespective of sex, and high-intensity smoking remained stable throughout all age groups (**Supplementary** Figure 2).

**Figure 1.**
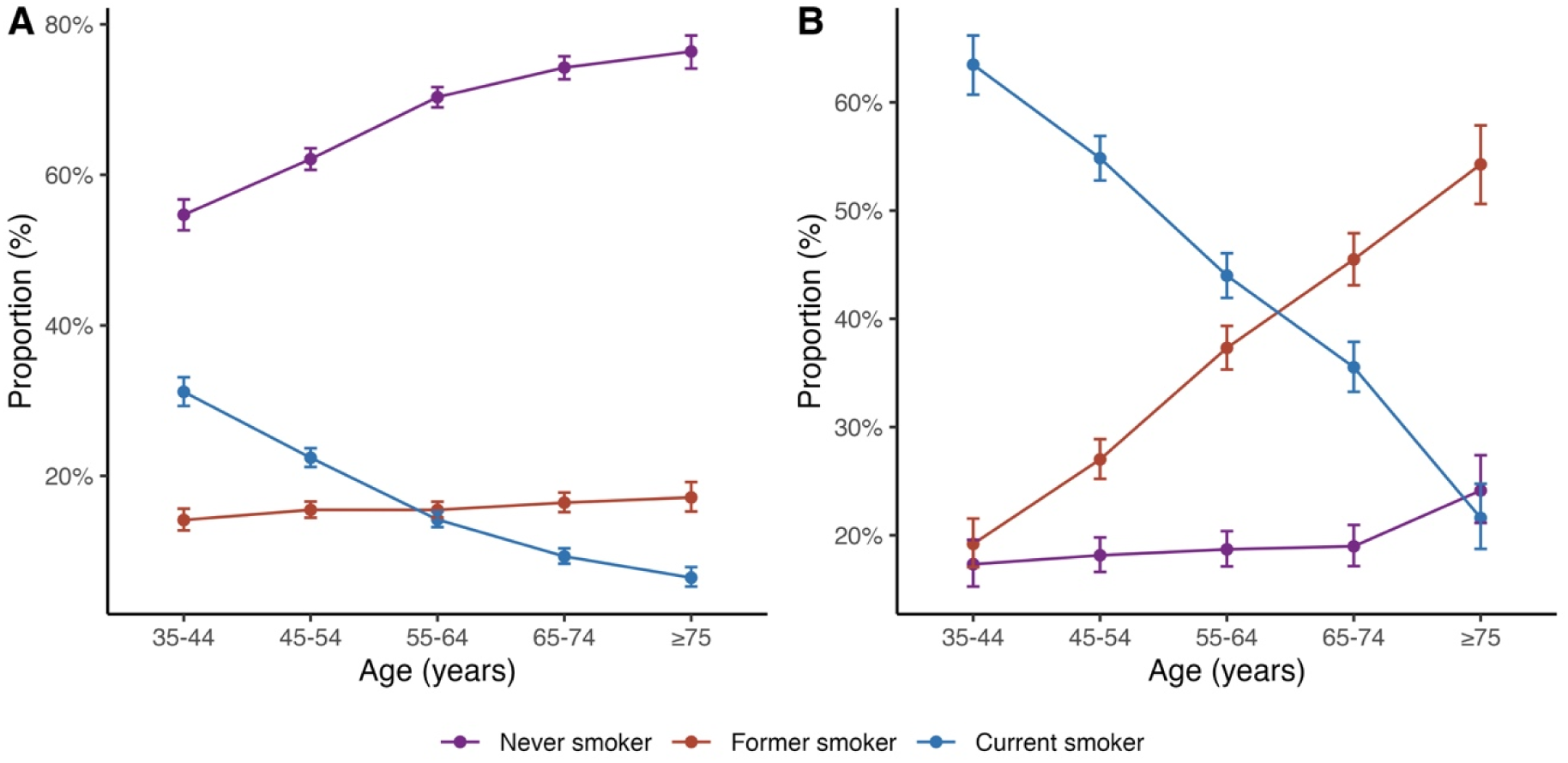
Smoking prevalence by age and sex. (A) Smoking status by age in women, (B) Smoking status by age in men.

Body mass index.

|  |  |  |  |  |  |  |
| --- | --- | --- | --- | --- | --- | --- |
| 3.02 (±11.19) | 52.65 (±10.35) | 58.62 (±12.3) | 61.45 (±11.67) | 52.98 (±10.94) | 55.38 (±11.21) | 54.62 (±10.45) |
| 30 (33.23%) | 123 (32.98%) | 514 (33.08%) | 1026 (35.69%) | 389 (33.33%) | 629 (38.17%) | 383 (39.04%) |
| 94 (7.26%) | 35 (9.38%) | 254 (16.34%) | 410 (14.26%) | 193 (16.54%) | 217 (13.17%) | 184 (18.76%) |
| 85 (22.02%) | 87 (23.32%) | 323 (20.79%) | 528 (18.37%) | 272 (23.31%) | 397 (24.09%) | 225 (22.94%) |
| 16 (55.33%) | 211 (56.57%) | 774 (49.81%) | 1521 (52.9%) | 608 (52.1%) | 858 (52.06%) | 460 (46.89%) |
| 99 (15.38%) | 40 (10.72%) | 203 (13.06%) | 416 (14.47%) | 94 (8.05%) | 176 (10.68%) | 112 (11.42%) |
| 20 (16, 29) | 18 (15, 24) | – | 16 (14, 20) | 18 (15, 20) | 17 (15, 20) | 16 (14, 19) |
| 3 (2, 5) | 15 (10, 20) | – | 5 (2, 20) | 2 (1, 3) | 4 (2, 6) | 15 (10, 20) |
| 74 (7, 10.84) | 8.37 (6.82, 10.93) | 8.46 (6.82, 10.48) | 8.1 (6.73, 10.29) | 8.83 (7, 10.75) | 8.65 (6.91, 10.75) | 9.01 (7, 10.84) |
| 23 (24.96%) | 112 (30.03%) | 442 (28.44%) | 914 (31.79%) | 287 (24.59%) | 435 (26.4%) | 236 (24.06%) |
| 10 (39.41%) | 135 (36.19%) | 626 (40.28%) | 1157 (40.24%) | 461 (39.5%) | 639 (38.77%) | 386 (39.35%) |
| 61 (35.63%) | 126 (33.78%) | 486 (31.27%) | 804 (27.97%) | 419 (35.9%) | 574 (34.83%) | 359 (36.6%) |
| 73.34 (±14.79) | 72.53 (±14.97) | 75.22 (±13.41) | 75.38 (±13.22) | 76.52 (±13.95) | 74.98 (±13.21) | 76.17 (±14.24) |
| 51.85 (±6.16) | 153.19 (±6.59) | 162.84 (±6.85) | 163.07 (±6.93) | 163.44 (±7) | 163.54 (±6.81) | 165.42 (±7.26) |
| 0.46 (±5.89) | 30.9 (±6.17) | 28.34 (±4.58) | 28.32 (±4.57) | 28.61 (±4.9) | 28 (±4.51) | 27.84 (±5.28) |
| 1.42 (±18.33) | 131.27 (±17.43) | 133.75 (±17.74) | 134.85 (±18.38) | 131.63 (±17.33) | 132.2 (±17.43) | 131.46 (±17.6) |
| 3.73 (±10.47) | 83.56 (±11.16) | 86.12 (±11.08) | 86.11 (±11.17) | 85.48 (±10.89) | 85.41 (±11.15) | 84.99 (±11.01) |
| 81 (13.99%) | 44 (11.8%) | 195 (12.55%) | 373 (12.97%) | 130 (11.14%) | 176 (10.68%) | 114 (11.62%) |
| 05 (46.75%) | 169 (45.31%) | 729 (46.91%) | 1501 (52.21%) | 541 (46.36%) | 788 (47.82%) | 453 (46.18%) |
| 80 (6.18%) | 21 (5.63%) | 74 (4.76%) | 115 (4%) | 44 (3.77%) | 63 (3.82%) | 34 (3.47%) |

### Smoking status of participants with diabetes at resurvey

Smoking status changed significantly during follow-up. Among 10,143 participants resurveyed, we identified 1,246 individuals initially classified with diabetes (diagnosed and undiagnosed) whose data was used in the main mortality analyses. Of these surviving participants, 666 (53.5%) were never smokers, 239 (19.2%) were former smokers and 341 (27.4%) were current smokers at baseline. Among never smokers, most remained as such and <1% started smoking during follow-up. Similarly, among former smokers, 71.5% remained as former smokers and only 3.7% restarted smoking (with the remaining 24.7% reporting never smoking at resurvey). However, changes in smoking status were most noticeable in current smokers. Over 59% reported quitting during the follow-up period, and only 28.4% continued smoking at resurvey. The median number of cigarettes smoked per day at resurvey among current smokers and former smokers was similar. At resurvey, those who were former smokers had better glycaemic control than current smokers (**Table 2, Supplementary** Figure 3).

**Table 2.**
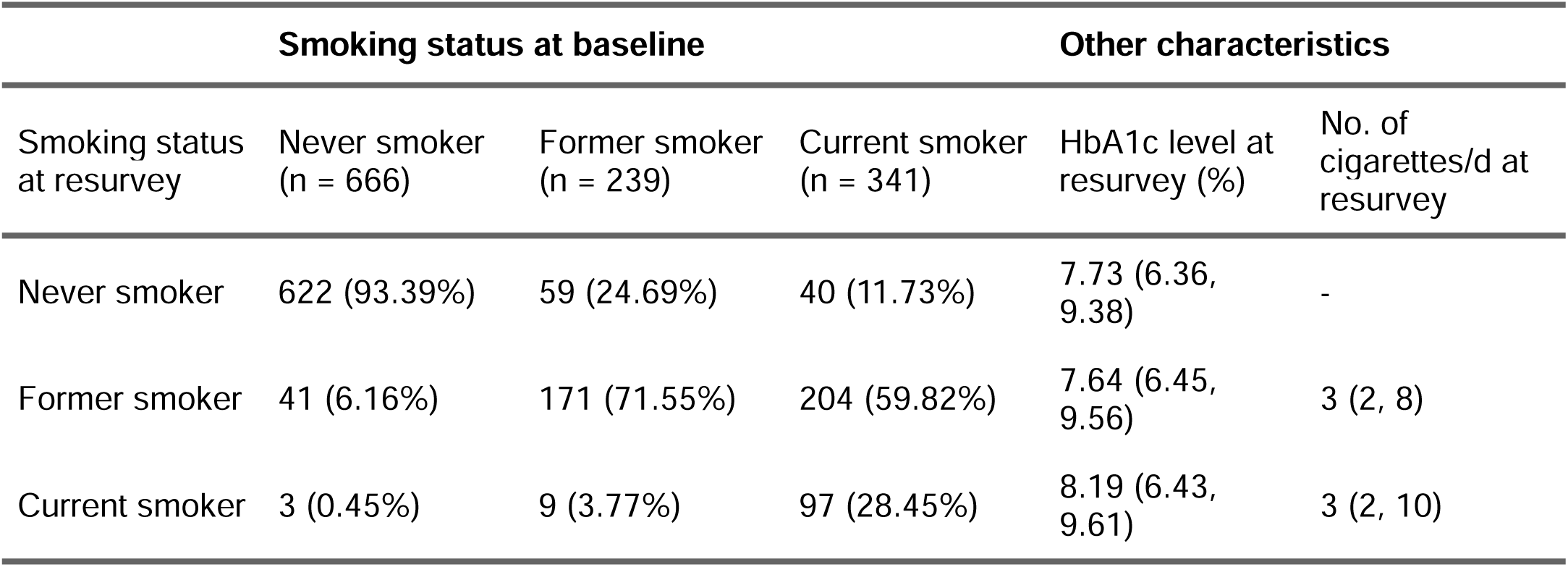
Changes in reported smoking habits from baseline to resurvey. HbA1c: Glycated haemoglobin.

### All-cause mortality and smoking status among individuals with diabetes

Among 24,200 participants with diabetes and after a median follow-up of 17.65 years, there were a total of 5,087 deaths at ages 35 to 74 years, 2,893 in women and 2,194 in men. Compared to never smokers, there was an increased risk for all-cause mortality among former (HR 1.08, 95%CI 1.01–1.17) and current smokers (HR 1.11, 95%CI 1.03–1.20, **Figure 2A**), with similar relative risks in daily and non-daily smokers (**Figure 2B**). Among daily smokers, there was increased risk in individuals who smoked ≥10 cigarettes/day (HR 1.28, 95%CI 1.14–1.43, **Figure 2C**). There was similar mortality risk across age categories, being highest among those who started smoking <15 years (HR 1.19, 95%CI 1.03–1.37, **Figure 2D**). Exploring all-cause mortality risk in former smokers, we found increased risk for participants who previously smoked ≥10 cigarettes/day (HR 1.16, 95%CI 1.02–1.31) but similar RRs irrespective of age starting (**Figure 2E, F**). Estimating attributable fraction, our results indicate that 7% of deaths in former smokers and around 22% in ≥10 cigarettes/day smokers aged 35-74 years could be prevented by avoiding smoking exposure in persons with diabetes.

**Figure 2.**
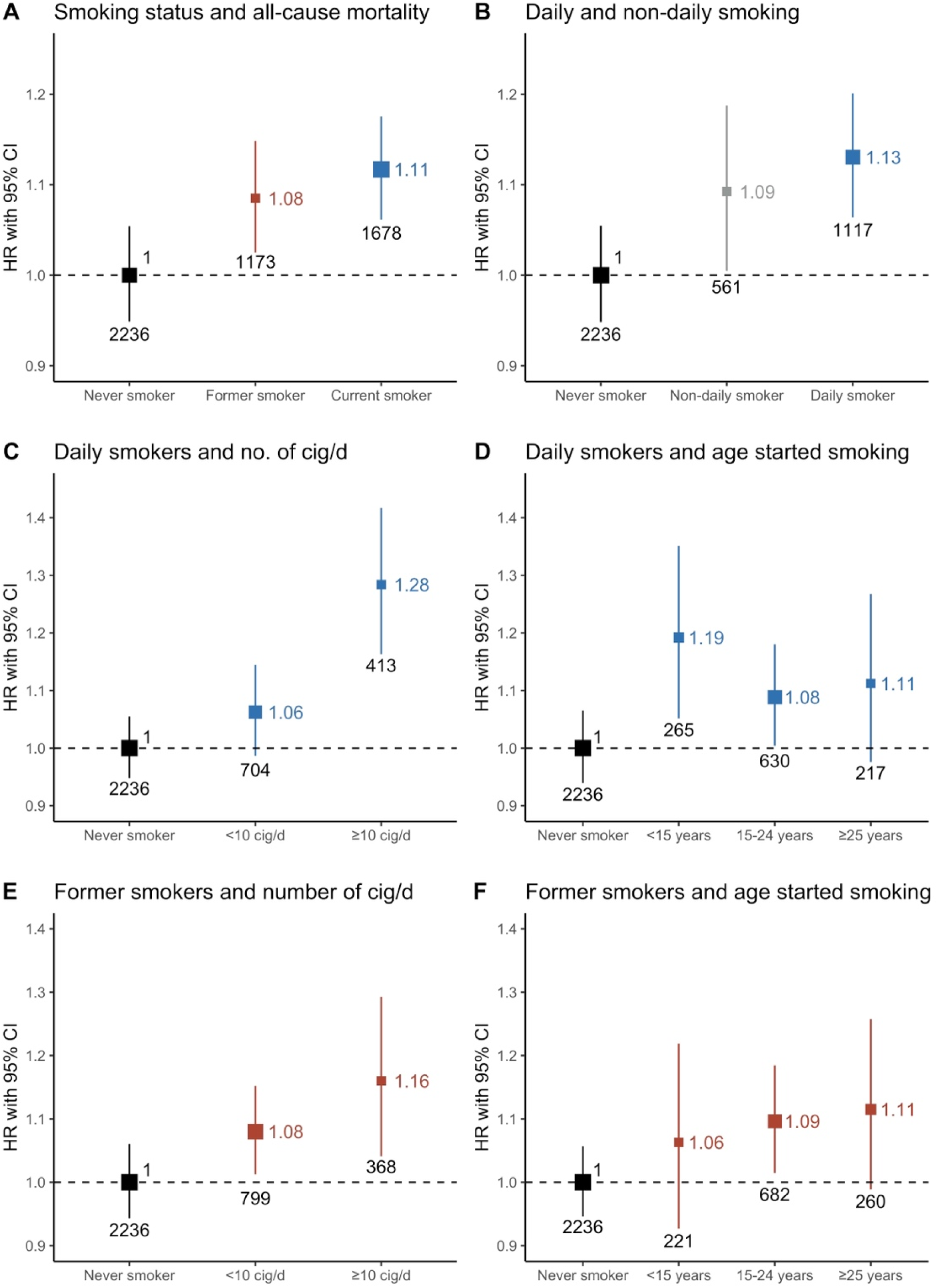
Hazard ratios for all-cause mortality by smoking status in individuals with diabetes. (A) Smoking status and all-cause mortality, (B) Non-daily, daily smoking and all-cause mortality in current smokers, (C) Number of cigarettes/day and all-cause mortality in daily smokers, (D) Age started smoking and all-cause mortality in daily smokers, (E) Number of cigarettes/day and all-cause mortality in former smokers, (F) Age started smoking and all-cause mortality in former smokers. All models were stratified by sex and age at risk (5-year age groups) and adjusted for place of residence (Coyoacán or Iztapalapa), educational level (elementary, high school, university, other), body mass index (BMI), HbA1c, time since diabetes diagnosis, and alcohol consumption.

### Smoking and cause-specific mortality among individuals with diabetes

Compared to never smokers, daily smokers had a significantly higher risk of cardiovascular mortality especially in those smoking ≥10 cigarettes/day (**Figure 3**). There was an increased risk of myocardial infarction related mortality in participants who smoked ≥10 cigarettes/day, higher risk of other vascular deaths in those who smoked ≥10 cigarettes/day, and a significantly higher risk for acute diabetes complications in participants who smoked ≥10 cigarettes/day. As expected, there were higher risks of death due to lung cancer and COPD especially in those who smoked <10 and ≥10 cigarettes per day, respectively. There was no association between smoking and mortality due to renal causes, stroke, hepatobiliary or gastrointestinal deaths, nor non-lung cancer, although the power to detect an association was probably low in the latter four categories. Finally, among former smokers, there was an increased risk of death due to acute diabetes complications, COPD, and hepatobiliary (**Supplementary** Figure 4).

**Figure 3.**
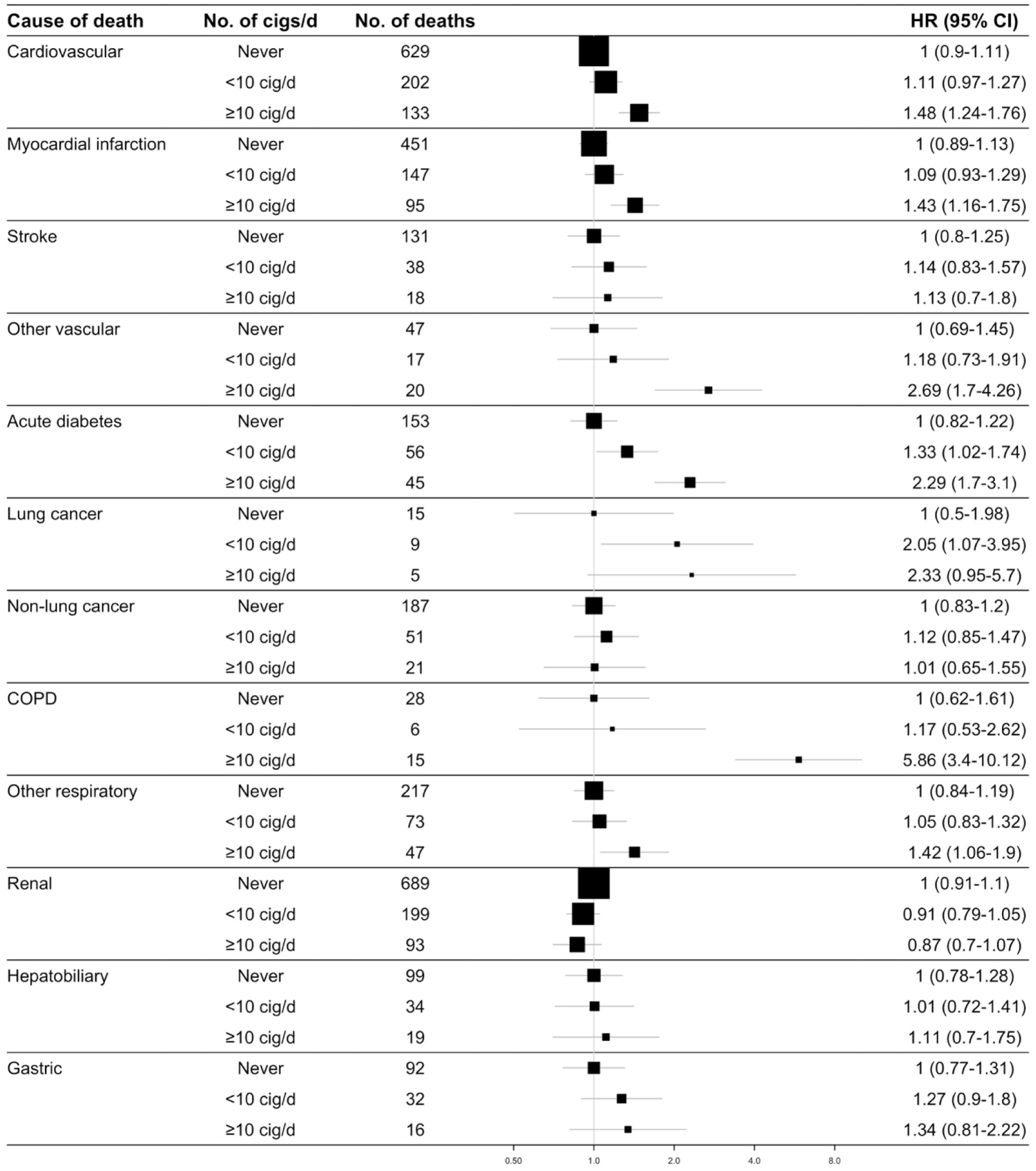
Cause-specific mortality in individuals with diabetes according to number of cigarettes smoked per day amongst daily smokers. COPD: Chronic obstructive pulmonary disease. All models were stratified by sex and age at risk (5-year age groups) and adjusted for place of residence (Coyoacán or Iztapalapa), educational level (elementary, high school, university, other), body mass index (BMI), HbA1c, time since diabetes diagnosis, and alcohol consumption.

### Sensitivity analysis

Separate models for men and women were fitted to determine whether there are different risks by sex, results which are shown in **Supplementary** Figure 5. There were a total of 3,528 deaths at ages ≥75 years, 2,198 in women and 1,330 in men. The estimated RR are shown in **Supplementary** Figure 6.

## DISCUSSION

In this study of 24,200 participants with diabetes and no other comorbidities in the Mexico City Prospective Study, daily and non-daily smoking were associated with an increased risk for all-cause mortality in a dose-dependent manner. Daily smokers had higher mortality risk in those who smoked ≥10 cigarettes/day and in participants who started smoking aged <15 years. Notably, over 50% of current smokers at baseline had stopped smoking at resurvey, which suggests a significant change in smoking habits during follow-up. Daily smokers were also at increased risk of death due to causes directly related to smoking (cardiovascular disease, myocardial infarction, other vascular disease, lung cancer and COPD), but also due to acute diabetes complications. Among former smokers with diabetes, a significant proportion remained as such at resurvey, with increased mortality risk in those who previously smoked ≥10 cigarettes/day and in those who started smoking at ages 15-24. Former smokers were at increased risk of death primarily due to COPD, hepatobiliary disease and, interestingly, acute diabetes complications as well. These findings confirm that smoking represents an excess risk that is modifiable in this population and support the notion that it remains a significant risk factor to be considered to reduce cardiovascular risk as part of long-term diabetes management.

The effect of smoking on all-cause and cause-specific mortality has been extensively reported in the general population^14,15^ and numerous studies have also explored the association between smoking and mortality risk in individuals with diabetes, finding a predominately positive association^16–18^. Regarding cause-specific mortality, smoking among individuals with diabetes has been associated with significantly increased risk for coronary heart disease, stroke, and peripheral arterial disease^19,20^. Our findings in this subgroup of participants are in line to those reported previously for all-cause mortality, albeit with lower estimates for both former and daily smokers^19^; and our cause-specific mortality analysis identified similar risk associations, primarily with cardiovascular disease and myocardial infarction for heavy smokers, but not with stroke probably related to lack of statistical power in our study population. Interestingly, our results show a significant impact on death related to acute hyperglycaemic crises in former and daily smokers. Smoking is not considered a typical risk factor for the development of either diabetic ketoacidosis or hyperglycaemic hyperosmolar state; however concomitant illness such as myocardial infarction or cerebrovascular events can be precipitating causes (among others like poor adherence to treatment or infection)^21,22^. Consequently, the observed association in our analysis might be due to poor baseline glycaemic control and the known association between smoking and cardiovascular events, which might lead to higher risk of developing a precipitating cause and increased risk of acute glycaemic crises mortality in daily and former smokers. These findings should further strengthen the fact that smoking cessation represents a pivotal intervention to reduce mortality risk for individuals with diabetes.

In Mexico, a previous analysis of the MCPS on healthy individuals found that smoking was responsible for significantly higher mortality risk among active smokers with a dose-response relationship^6^, representing the first prospective analysis of smoking-related mortality in Mexican population. Although smoking prevalence in Mexico declined significantly from 2000 to 2010, there has been a slight increase in prevalence between 2011 and 2016. Moreover, prevalence of previously diagnosed diabetes has increased dramatically in Mexico reaching 10.2% in 2021^3^. Even though smoking cessation is advised for every person with diabetes, it has been suggested that individuals with chronic diseases, particularly younger persons, have higher prevalence of smoking^23,24^. Notably, the smoking trends among individuals with diabetes in Mexico are largely unknown, and so is the mortality risk associated with smoking, thus the rational to explore tobacco use in this subgroup of participants. It should be noted as well that previous analyses of MCPS have found high mortality rates in individuals with diabetes, which accounts for at least a third of all deaths at ages 35-74 years^25^. Given this conditions, even if the smoking-related RRs found in this analysis are somewhat lower than previously reported in individuals with diabetes^19^ and in disease-free participants^6^, the smoking-related mortality risk for individuals with diabetes might be similar or even greater in this population. Finally, it should be highlighted that several interventions have proved effective to promote smoking cessation in individuals with diabetes^4,26^, which emphasizes the importance of improving public health policies directed towards smoking cessation.

### Strengths and limitations

Our study has several strengths, including its prospective design, which allows to establish a potentially causal association between mortality risk and smoking in individuals with diabetes, the large sample size obtained from two urban districts in Mexico City and the analysis of resurvey data to assess changes of smoking habits during follow-up. We recognize however some limitations which should be considered to adequately interpret our findings. First, information regarding characterization of former smokers is incomplete, which did not allow us to estimate the effect of time since smoking cessation in all-cause mortality. Second, because resurvey information was collected only in a subset of surviving participants, we were unable to explore changes in smoking exposure during follow-up for every individual and, consequently, our estimates using baseline interview information do not account for changes in smoking patterns over time, which might lead to a potential underestimation of RRs. Reverse causality might also be a concern. Diabetes diagnosis might change the smoking status of participants which might be of concern in former smokers and in participants with undiagnosed diabetes. Given that we have limited information regarding why and when former smokers quitted and the precise moment undiagnosed participants were diagnosed with diabetes, caution should be taken regarding our estimates in these categories. Finally, due to the observational nature of the study, we are unable to rule out residual confounding despite controlling for known modifiers of mortality risk in statistical analyses.

### Conclusions and perspectives

In summary, our results highlight that smoking in individuals with diabetes still represents an important risk factor for all-cause and cause-specific mortality. Former smokers with diabetes are at increased risk dependent on previous smoking intensity, whilst daily smokers have a dose-dependent increase in mortality risk, particularly for individuals who started smoking at younger ages. Notably, smoking is associated with higher risk of acute diabetes complications, which support the view that smoking cessation is still a relevant target to improve diabetes management and that it should be strongly recommended during patient counselling.

## ACKNOWLEDGMENTS

This project was registered and approved by the Research Committee at Instituto Nacional de Geriatría, project number DI-PI-006/2020. CAFM is enrolled at the PECEM Program of the Faculty of Medicine at UNAM. DRG and CAFM are supported by CONACyT. The authors thank the participants for their willingness to take part in this prospective study 20 years ago. This research was conducted using Mexico City Prospective Study (MCPS) data obtained through an open-access data request (application number 2022-012). We want to thank Jonathan Emberson and Diego Aguilar-Ramirez for their very thoughtful comments and methodological support on this manuscript.

## AUTHOR CONTRIBUTIONS

Establishing the cohort: JBC, PKM, JAD and RTC. Obtaining funding: JBC, PKM, JAD, RTC and OYBC. Data acquisition, analysis, or interpretation of data: DRG, OYBC, CAFM, PSC, ANL, MRBA, LFC, AVV, JPE, JPDS, AKG, PAV, PKM, RTC, JAD, JAS, NEAV. Drafting first version of manuscript: DRG, OYBC. Critical revision of the report for important intellectual content: All authors. All authors have seen and approved the final version and agreed to its publication. Each author contributed important intellectual content during manuscript drafting or revision and accepts accountability for the overall work by ensuring that questions pertaining to the accuracy or integrity of any portion of the work are appropriately investigated and resolved.

## DATA AVAILABILITY

Data from the Mexico City Prospective Study are available to bona fide researchers. For more details, the study’s Data and Sample Sharing policy may be downloaded (in English or Spanish) from https://www.ctsu.ox.ac.uk/research/mcps. Available study data can be examined in detail through the study’s Data Showcase, available at https://datashare.ndph.ox.ac.uk/mexico/. Code is available for reproducibility of results at https://github.com/oyaxbell/smoking_diabetes_mcps/

