## Supplementary Material for "Smoking, all-cause, and cause-specific mortality in individuals with diabetes in Mexico: an analysis of the Mexico City Prospective Study"

**SUPPLEMENTARY FIGURES**

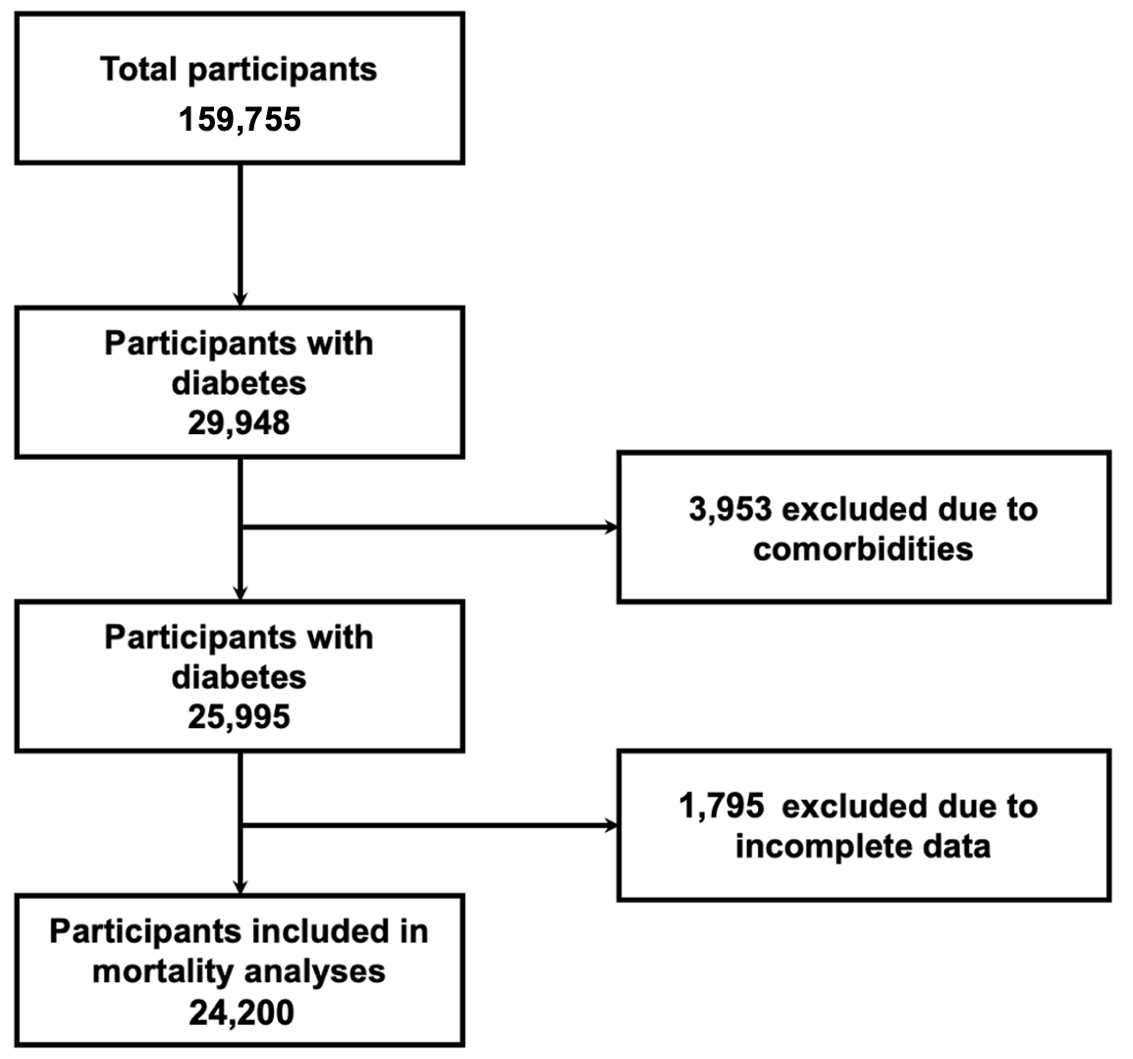

**Supplementary Figure 1.** Flowchart of study participants included in mortality analyses.

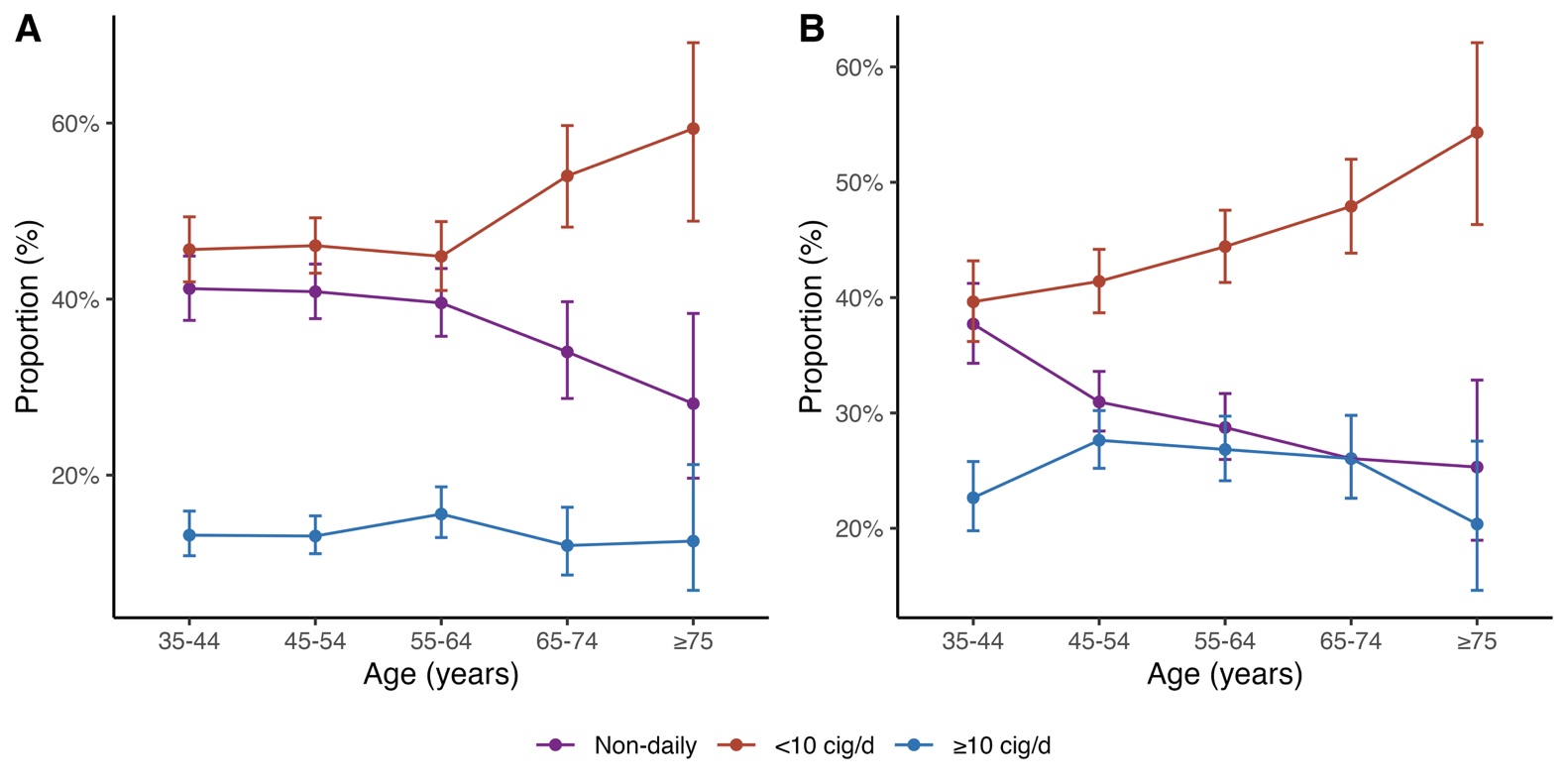
**Supplementary Figure 2.** Smoking prevalence by age and sex amongst current smokers. (A) Smoking status by age in women, (B) Smoking status by age in men.

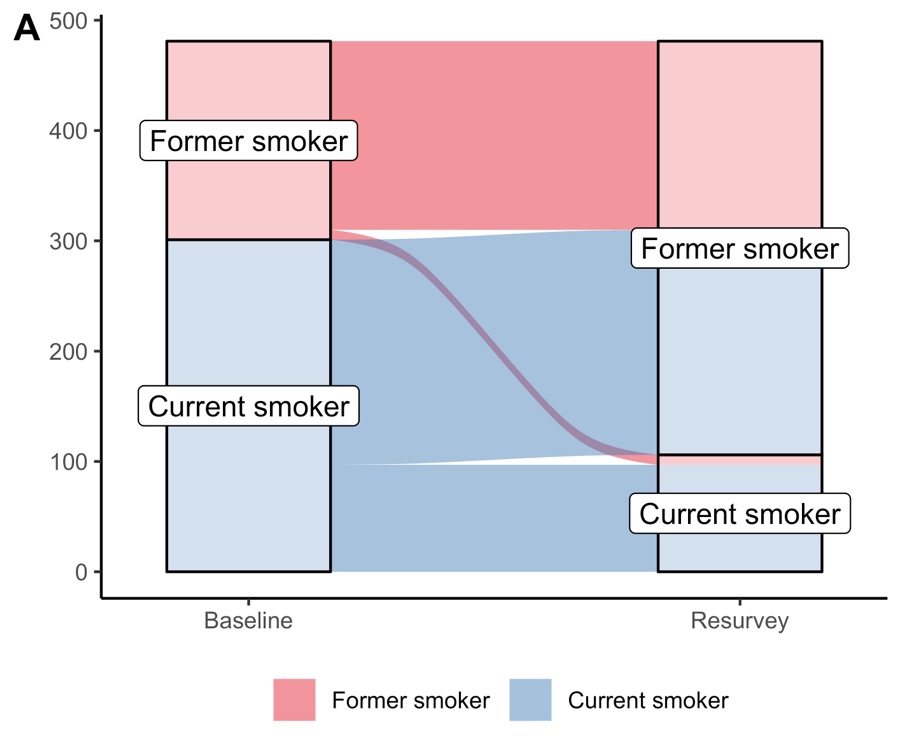

**Supplementary Figure 3.** (A) Changes in smoking status from baseline interview to resurvey in 1,246 individuals with diabetes.

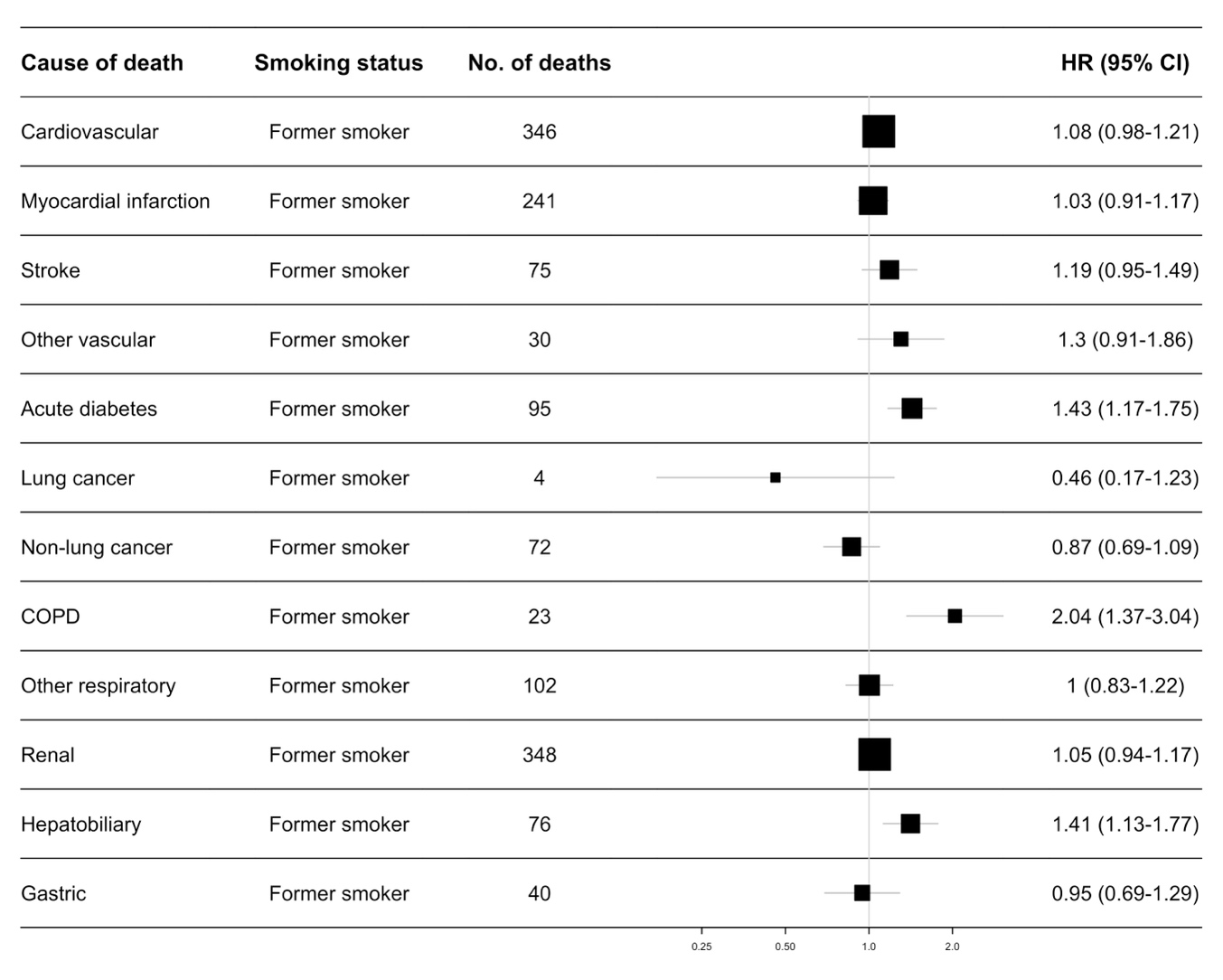

**Supplementary Figure 4.** Cause-specific mortality in individuals with diabetes amongst former smokers using never smokers as comparison. COPD: Chronic obstructive pulmonary disease. All models were stratified by sex and age at risk (5-year age groups) and adjusted for place of residence (Coyoacán or Iztapalapa), educational level (elementary, high school, university, other), body mass index (BMI), HbA1c, time since diabetes diagnosis, and alcohol consumption.

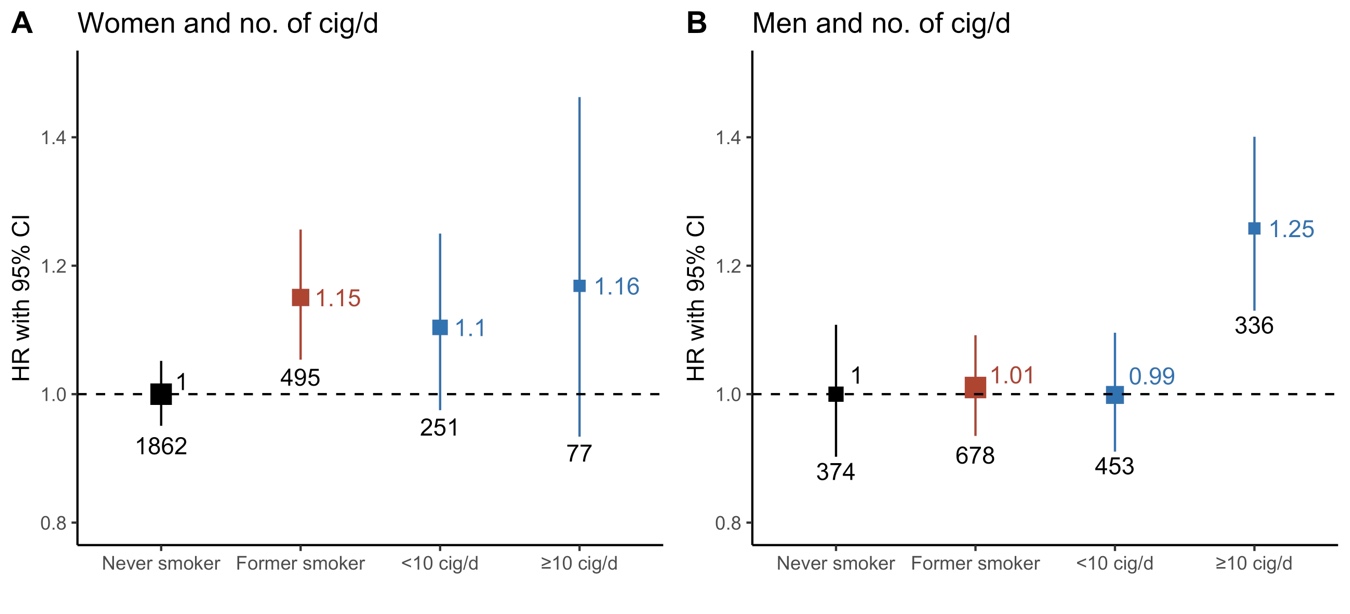

**Supplementary Figure 5.** Separate Cox models for (A) women and (B) men to estimate adjusted HR according to smoking status and number of cigarettes/day. All models were stratified by sex and age at risk (5-year age groups) and adjusted for place of residence (Coyoacán or Iztapalapa), educational level (elementary, high school, university, other), body mass index (BMI), HbA1c, time since diabetes diagnosis, and alcohol consumption.

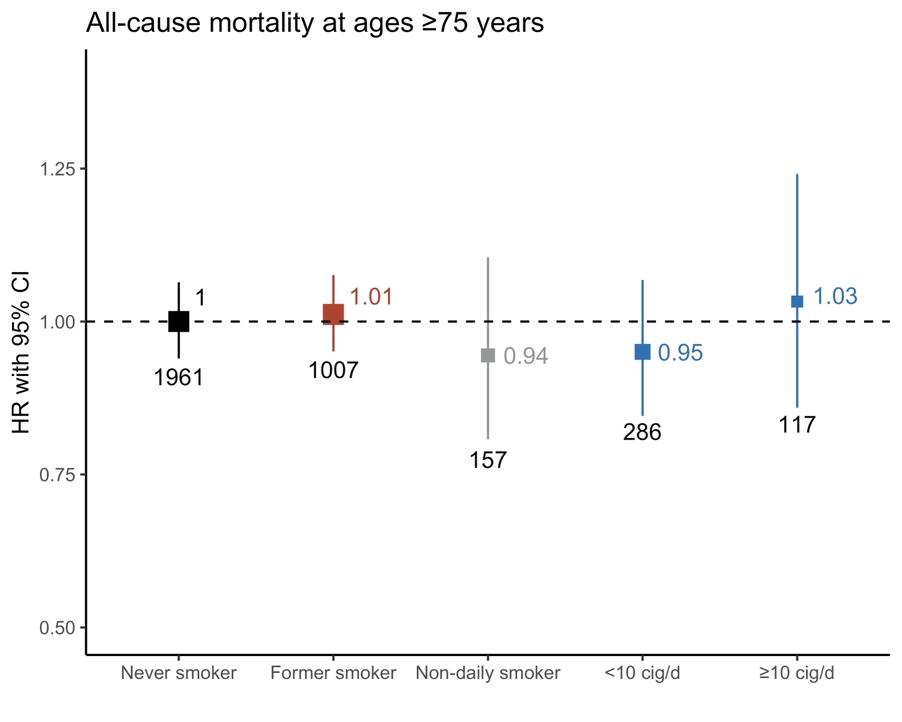

**Supplementary Figure 6.** Hazard ratios for all-cause mortality by smoking status in individuals with diabetes with deaths at ages ≥75 years. The model was stratified by sex and age at risk (5-year age groups) and adjusted for place of residence (Coyoacán or Iztapalapa), educational level (elementary, high school, university, other), body mass index (BMI), HbA1c, time since diabetes diagnosis, and alcohol consumption.

**SUPPLEMENTARY TABLES**

| **Cause of death** | **ICD-10 codes (No. of deaths)** | **Total number of deaths** |
| --- | --- | --- |
| Cardiovascular | E115 (31), E145 (30), I059 (4), I099 (2), I110 (36), I119 (2), I200 (2), I209 (1), I210 (16), I211 (2), I219 (777), I249 (9), I251 (19), I252 (1), I258 (4), I259 (38), I269 (19), I272 (1), I279 (4), I319 (1), I330 (3), I350 (4), I38X (3), I420 (2), I442 (5), I443 (1), I460 (1), I469 (7), I472 (1), I489 (1), I490 (2), I499 (2), I500 (16), I501 (4), I509 (32), I515 (1), I518 (1), I519 (4), I608 (1), I609 (17), I61 (1), I612 (1), I614 (1), I615 (1), I619 (80), I620 (2), I629 (1), I633 (3), I634 (5), I635 (2), I638 (1), I639 (26), I64X (45), I669 (4), I672 (1), I674 (2), I678 (35), I679 (51), I693 (4), I694 (2), I698 (13), I710 (1), I714 (1), I718 (2), I739 (1), I771 (11), I802 (2), I803 (1), I830 (1), I99X (1), K550 (21), K551 (1), K552 (1), K559 (1), R570 (8) | 1441 |
| Cardiac | I059 (4), I099 (2), I110 (36), I119 (2), I200 (2), I209 (1), I210 (16), I211 (2), I219 (777), I249 (9), I251 (19), I252 (1), I258 (4), I259 (38), I272 (1), I279 (4), I319 (1), I330 (3), I350 (4), I38X (3), I420 (2), I442 (5), I443 (1), I460 (1), I469 (7), I472 (1), I489 (1), I490 (2), I499 (2), I500 (16), I501 (4), I509 (32), I515 (1), I518 (1), I519 (4), R570 (8) | 1017 |
| Stroke | I608 (1), I609 (17), I61 (1), I612 (1), I614 (1), I615 (1), I619 (80), I620 (2), I629 (1), I633 (3), I634 (5), I635 (2), I638 (1), I639 (26), I64X (45), I669 (4), I672 (1), I674 (2), I678 (35), I679 (51), I693 (4), I694 (2), I698 (13) | 299 |
| Other vascular | E115 (31), E145 (30), I269 (19), I710 (1), I714 (1), I718 (2), I739 (1), I771 (11), I802 (2), I803 (1), I830 (1), I99X (1), K550 (21), K551 (1), K552 (1), K559 (1) | 125 |
| Acute diabetes | E100 (3), E101 (3), E110 (125), E111 (112), E140 (92), E141 (69), E162 (5) | 409 |
| Lung cancer | C029 (1), C142 (1), C329 (2), C349 (33), D380 (1) | 38 |
| Non-lung cancer | C159 (4), C169 (44), C170 (2), C182 (1), C184 (1), C189 (30), C20X (7), C220 (10), C221 (8), C229 (30), C23X (10), C240 (4), C241 (3), C249 (4), C250 (6), C259 (20), C260 (2), C269 (1), C412 (1), C419 (1), C438 (1), C439 (2), C457 (1), C480 (3), C482 (1), C492 (1), C499 (1), C509 (25), C519 (1), C539 (23), C541 (3), C55X (1), C56X (14), C609 (1), C61X (8), C64X (15), C66X (1), C679 (6), C709 (1), C710 (3), C719 (3), C73X (6), C741 (1), C759 (1), C760 (1), C762 (1), C763 (2), C765 (1), C780 (1), C786 (2), C787 (2), C800 (4), C809 (1), C819 (2), C829 (1), C833 (1), C838 (1), C839 (1), C859 (7), C901 (1), C910 (2), C920 (6), C921 (2), C959 (2), D371 (1), D374 (1), D376 (2), D410 (1), D419 (1), D430 (3), D432 (1), D487 (1), D489 (1) | 365 |
| COPD | J42X (5), J439 (6), J440 (30), J448 (2), J449 (35) | 78 |
| Other respiratory | A162 (2), A165 (1), A169 (2), B441 (1), J069 (1), J09 (1), J111 (1), J151 (1), J159 (14), J180 (25), J181 (15), J182 (2), J188 (1), J189 (281), J209 (1), J22X (11), J391 (1), J459 (4), J46X (2), J60X (1), J677 (1), J690 (1), J81X (2), J841 (14), J849 (1), J850 (1), J869 (5), J90X (4), J939 (1), J949 (1), J960 (2), J961 (1), J969 (4), J981 (1), J984 (3), J985 (2), J988 (1), U071 (47), U072 (44) | 504 |
| Renal | E102 (10), E112 (752), E122 (1), E142 (293), I120 (43), I130 (1), I131 (1), I132 (17), N009 (2), N039 (18), N059 (2), N10X (2), N119 (1), N12X (4), N142 (1), N151 (6), N179 (36), N180 (4), N185 (21), N189 (119), N19X (43), N200 (2), N289 (1), N390 (127), Y841 (1) | 1508 |
| Hepatobiliary | B171 (6), B181 (1), B182 (1), I850 (5), K563 (1), K701 (1), K703 (28), K704 (4), K709 (4), K711 (1), K720 (2), K721 (14), K729 (40), K739 (2), K746 (78), K750 (4), K759 (1), K766 (4), K767 (4), K769 (6), K800 (1), K801 (3), K802 (2), K803 (2), K804 (1), K805 (2), K810 (5), K819 (3), K822 (1), K829 (1), K830 (6), K831 (1), K852 (1), K858 (6), K859 (12), K85X (6), K861 (2), K868 (4) | 266 |
| Gastric | A047 (1), A060 (1), A090 (9), A099 (9), A09X (8), K102 (1), K222 (1), K254 (3), K255 (5), K259 (2), K264 (1), K274 (1), K290 (2), K291 (1), K318 (3), K353 (1), K358 (2), K359 (2), K419 (1), K430 (2), K440 (1), K460 (1), K461 (1), K469 (2), K529 (1), K562 (1), K564 (1), K566 (10), K572 (1), K578 (1), K579 (3), K610 (2), K612 (1), K630 (1), K631 (7), K632 (2), K635 (1), K639 (1), K650 (8), K658 (1), K659 (31), K669 (1), K920 (8), K922 (54) | 197 |

**Supplementary Table 1.** ICD-10 codes according to cause of death with number of events in parenthesis.
